# Short-term high-dose nicotinamide treatment across glaucoma subtypes reveals increased mtDNA content and minimal metabolomic change in blood

**DOI:** 10.1101/2025.07.18.25331757

**Authors:** Antoni Vallbona-Garcia, James R Tribble, Simon T. Gustavsson, Birke J Benedikter, Patrick J Lindsey, Carroll AB Webers, Hubert JM Smeets, Gauti Jóhannesson, Theo GMF Gorgels, Pete A Williams

## Abstract

Nicotinamide (NAM) supplementation has emerged as a potential treatment for the retinal ganglion cell (RGC) degeneration of the eye disorder glaucoma. In glaucoma, RGC degeneration has been linked to declining mitochondrial metabolic capacity. NAM is a precursor for the synthesis of nicotinamide adenine dinucleotide (NAD) through the salvage pathway. NAD is a REDOX cofactor and an essential metabolite for cellular functions and the production of energy through mitochondria. In this study, we assessed the effects of a 2- week NAM supplementation treatment on blood NAM levels, NAM metabolism, and wider blood metabolome in 90 glaucoma subjects from 3 different glaucoma subtypes (high tension glaucoma (HTG), normal tension glaucoma (NTG), and pseudoexfoliative glaucoma (PEXG), n = 30 per group), and 30 age and sex-matched healthy controls. We performed small-molecular-weight high-resolution mass spectrometry to assess the metabolome and qPCR to assess the mtDNA amount per cell. At baseline (pre-NAM), only ethylmalonic acid, a compound related to defects in β-oxidation and mitochondrial dysfunction, was found to be modestly increased in the 3 glaucoma subtypes in comparison to controls. All groups showed a similar metabolome response to treatment with a specific increase in NAM and related species (1-methylnicotinamide, 6-hydroxynicotinamide, N1-methyl-2-pyridone-5- carboxamide), an increase in 5-methylcytosine, and a decrease in 4-pyridoxic acid. A linear model corrected by age and sex demonstrated that between groups, only Sarcosine had a different response, with a small reduction in HTG and NTG post-treatment. NAM treatment resulted in a significant but slight within-group increase in blood mtDNA amount in controls and HTG (∼12% and ∼17%, respectively), as determined by a generalized linear mixed effects model adjusted for age and sex. This study suggests that NAM treatment leads to similar plasma metabolome changes between glaucoma groups and controls, which predominantly reflect increased NAM metabolites and intermediates, with minimal effects on the wider metabolome, and a modest increase in mtDNA amount in HTG and controls. As this was observed in a short-term accelerated dosing context, long-term studies will be required to provide more information on the long-term effects of oral NAM supplementation.

## 1. Introduction

Glaucoma is the leading cause of irreversible blindness in the world, estimated to affect more than 70 million people.^1^ It encompasses a group of optic neuropathies characterized by neurodegeneration of retinal ganglion cells (RGCs, whose axons form the optic nerve).^2–4^ The main risk factor of glaucoma is elevated intraocular pressure (IOP), but several other factors have also been found associated with its pathophysiology, e.g., ageing and variants in over 50 genes.^5–7^ Elevated IOP is the only modifiable risk factor and the target for current glaucoma treatments either through medication, laser therapy or surgery, aimed to slow down or stop disease progression.^8–10^ However, reducing IOP does not prevent disease progression in all cases, and a considerable proportion of glaucoma patients eventually go blind during their lifetime.^11,12^ This underscores the need to develop additional, complementary treatment options. Currently, there are no therapies that specifically target RGCs.^5,13^

In recent years, mitochondria and the RGCs’ bioenergetic risk have emerged as promising targets. Mitochondria are the key cell energy organelles, producing ATP through a group of protein complexes (oxidative phosphorylation; OXPHOS). Mitochondria have been proposed as a disease component and neuroprotective target in many neurodegenerative diseases.^14–18^ In the case of glaucoma, mutations in both the nuclear mitochondrial encoded genes (*OPA1, MFN1, MFN2, OPTN*), and in the mitochondrial genome (mtDNA) have been linked to the disease as risk factors.^7,19–23^ Functional mitochondrial impairments have been characterized at a systemic level in blood cells of glaucoma subjects.^24–26^ mtDNA damage (deletions, mutations) and mitochondrial structural defects have been observed in RGCs, retina, and other eye structures of glaucoma patients and animal models.^27–30^ RGCs are enriched with mitochondria and have a high energy demand. As such, it is likely that suboptimal mitochondrial function can pose a bioenergetic risk that predisposes to or exacerbates neurodegeneration in combination with an age-related decline.^31–34^

Conversely, improving the bioenergetic health of RGCs may represent a neuroprotective treatment for glaucoma. This hypothesis is currently tested by nicotinamide (NAM) supplementation, aiming to increase NAD content.^35–37^ NAM is a precursor of NAD via the salvage pathway. NAD+, the oxidized form, is reduced to NADH in several biological pathways, e.g., glycolysis, beta-oxidation of fatty acids, and citric acid cycle.^38,39^ NADH acts as the first electron donor for the mitochondrial ATP production through the OXPHOS system. In animal models of glaucoma, NAD declines in the retina and optic nerve. ^35^ Supplementation with NAM (or gene therapies increasing salvage pathway enzymes to drive increased NAD production) has been demonstrated to be neuroprotective, preventing mitochondrial and metabolic dysfunction as well as RGC degeneration.^35,40,41^ NAM levels in the plasma of POAG patients have been demonstrated to be lower than in controls.^42,43^ NAD salvage pathway enzymes are reduced in the retina and optic nerve in retinas from glaucoma patients.^44^ Short-term clinical trials using NAM in glaucoma patients have demonstrated improvement in the inner retinal function after 12 weeks of treatment,^36^ and improvements in visual function in combination with pyruvate, ^45^ providing encouragement for longer clinical trials that are underway (NCT05275738). NAM is readily bioavailable to the central nervous system (CNS) and can alter the retinal metabolome of rodents within days.^35^ We previously identified effects after 2 weeks of NAM treatment, where NAM improved ocular blood supply in glaucoma patients, but the potential metabolic effects have yet to be explored in these patients.^46^

In the current study, we investigated the plasma metabolome and the buffy coat mtDNA content prior to and after an accelerated dosing strategy of 1.5 g/day of oral NAM in the first week and 3.0 g/day the second week. This was done in 3 different glaucoma subtypes, including high-tension glaucoma (HTG), normal-tension glaucoma (NTG), and pseudoexfoliative glaucoma (PEXG) and in age- and sex-matched healthy controls. The objectives of the study were:

- To explore the effects of a short-term NAM treatment on blood NAM levels, NAM metabolism, and the wider blood metabolome
- To study the effects of NAM treatment on a systemic mitochondrial biomarker, i.e. the mtDNA content of the blood cells.

## 2. Materials and methods

### 2.1 Ethical approval and patient sampling

The human studies adhered to the tenets of the Declaration of Helsinki, and the ethics protocols were approved by the Swedish Ethical Review Authority (2020-01525, 2021- 01036, 2021-03745, and 2022-04851).

Venous blood samples were collected from each study participant in EDTA tubes and serum tubes (the latter containing silicon dioxide for pro-coagulative effects) at baseline (pre-NAM) and after two weeks of nicotinamide treatment (post-NAM), at least three hours after the last meal. The tubes were sent immediately after sampling to a centralized biobank (Biobanken Norr) in the same hospital where the samples were collected (Umeå University Hospital). At the biobank, the tubes underwent centrifugation (1500G, 15 minutes, room temperature), and aliquots (all 250 µL) were collected from the tubes using a Hamilton Microlab Star ^LET^. Aliquots of erythrocytes, plasma, and buffy coat were collected from the EDTA tubes. From the serum tubes, aliquots of serum were collected. All the aliquots were placed in SBS format cryotubes and then immediately stored at −80°C until further analysis. The time between sampling and storage was 37 – 180 minutes (mean value 80 minutes, median value 77 minutes). The aliquots were kept frozen at −80°C as they were sent for analysis.

In the study, 90 glaucoma (30 HTG, 30 NTG and 30 PEXG) and 30 age- and sex-matched healthy controls were included. The mentioned blood fractions were collected and isolated pre- and post-2 weeks of NAM supplementation. Out of those 120 participants, the following samples were available for this study: 232 plasma samples (*n* = 30 controls, *n* = 30 HTG, *n* = 30 NTG, *n* = 28 PEXG; pre-NAM / *n* = 29 controls, *n* = 29 HTG, *n* = 29 NTG, *n* = 27 PEXG; 114post-NAM treatment) and 234 buffy coat samples (*n* = 30 controls, *n* = 30 HTG, *n* = 30 NTG, *n* = 29 PEXG; 119 pre-NAM / *n* = 30 controls, *n* = 27 HTG, *n* = 29 NTG, *n* = 29 PEXG; 115post-NAM). Some samples were not available due to the participant not being able to join the first or second study visit, or the blood fraction could not be properly collected and stored. In the case of plasma, samples were excluded if participants had not fasted for at least 3 hours before sample collection.

### 2.2 Metabolomics

#### 2.2.1 Sample processing

Sample preparation of plasma samples was performed with a semi-automated approach using an Agilent Bravo automatic liquid handling platform (G5563AA, Agilent Technologies, CA, USA) using the accompanying software Agilent VWorks for automation control. 720 µl of extraction buffer (90/10 v/v methanol: water) including internal standards was added to extract plates (96 Agilent 203426-100 PP, 1 mL Rnd Btm). 80 µl of the plasma samples were added to the extract plates and the plates were mixed using the Bravo system (at 1 000 rpm with 10 mixes). The proteins were precipitated at −20 °C for 2 hours. The sample plates were centrifuged at +4 °C, 4 000 rpm (3 488 g), for 10 minutes on a tabletop swingout rotor. 200µL supernatant was transferred to LC plates (96 Agilent WP 5043-9314 0.33 mL VBtm) using the Bravo system. The sample plates were evaporated to dryness under a stream of nitrogen (30 L min-1 at 30 °C) and stored at −80 °C until analysis.

#### 2.2.2 LC-MS HILIC Metabolomics

Before analysis the samples were re-suspended in 10 + 10 µL methanol and elution solvent A. The samples were analyzed in batches according to a randomized run order. Each batch of samples was first analyzed in positive mode. After all samples within a batch had been analyzed, the instrument was switched to negative mode and a second injection of each sample was performed.

The chromatographic separation was performed on an Agilent 1290 Infinity UHPLC-system (Agilent Technologies, Waldbronn, Germany). 1 μL of each sample were injected onto an Atlantis Premier BEH-Z-HILIC VanGuard FIT (1.7 µm, 2.1 x 50 mm) column (Waters Corporation, Milford, MA, USA) held at 40 °C. The HILIC gradient elution solvents were A) H_2_O, 10 mM ammonium formate, 5µM Medronic acid, pH 9, B) 90:10 Acetonitrile: H_2_O, 10 mM ammonium formate, pH 9. Chromatographic separation was achieved using a linear gradient (at a flow rate of 0.4 mL min^−1^) starting at 90% B decreasing to 80% B over 6 minutes; B was decreased to 20 % over 3.5 minutes and held at 20 % for 1.5 minutes; B was increased to 90 % for 0.5 minutes and the flow-rate was increased to 0.7 mL min^−1^ for 2 minutes; these conditions were held for 0.5 minutes, after which the flow-rate was reduced to 0.4 mL min^−1^ for 0.5 minutes before the next injection.

The compounds were detected with an Agilent 6546 Q-TOF mass spectrometer equipped with a jet stream electrospray ion source operating in positive or negative ion mode. A reference interface was connected for accurate mass measurements; the reference ions purine (4 μM) and HP-0921 (Hexakis(1H, 1H, 3H-tetrafluoropropoxy)phosphazine) (1 μM) were infused directly into the MS at a flow rate of 0.05 mL min^−1^ for internal calibration, and the monitored ions were purine m/z 121.05 and m/z 119.03632; HP-0921 m/z 922.0098 and m/z 966.000725 for positive and negative mode respectively. The gas temperature was set to 150°C, the drying gas flow to 8 L min^−1^ and the nebulizer pressure 35 psi. The sheath gas temp was set to 350°C and the sheath gas flow 11 L min^−1^. The capillary voltage was set to 4000 V in both positive and negative ion mode. The nozzle voltage was 300 V. The fragmentor voltage was 120 V, the skimmer 65 V and the OCT 1 RF Vpp 750 V. The collision energy was set to 0 V. The m/z range was 70 - 1700, and data was collected in centroid mode with an acquisition rate of 4 scans s^−1^. MSMS analysis was run on the QC samples (pooled metabolite extract from several samples) for identification purposes. Extraction blanks were included.

#### 2.2.3 Metabolomics analysis

All data pre-processing was performed using the Agilent MassHunter Profinder version B.10.0 SP1 (Agilent Technologies Inc., Santa Clara, CA, USA). The data pre-processing was performed in a targeted fashion. A pre-defined list of metabolites commonly found in plasma and serum were searched for using the Batch Targeted feature extraction in MassHunter Profinder. An-in-house LC-MS library built up by authentic standards run on the same system with the same chromatographic and mass-spec settings, were used for the targeted processing. Internal standards indicated RSD < 10% supporting good analytical quality. Batch correction was performed by normalizing to the Pooled QC (minus the extraction blank) for each batch. Statistical analysis was performed in MetaboAnalyst 5.0. All data were Pareto scaled. Baseline (pre-NAM) metabolomes were analyzed using a linear model with covariate adjustment for age and sex and *P* values were adjusted by false discovery rate (FDR). All subjects with a sample at baseline were included (*n* = 30 controls, *n* = 30 HTG, *n* = 30 NTG, *n* = 28 PEXG). Hierarchical clustering of pre- and post-NAM metabolites were performed by one minus *Pearson’s* correlation. Responses to NAM treatment by condition were analyzed by paired sample *Student’s t*-test and adjusted by FDR. Differences in response to NAM treatment were analyzed using a linear model with covariate adjustment for age, sex, time (pre/post NAM) and *P* values were adjusted by false discovery rate (FDR). Only subjects with both baseline and post-treatment samples were included (*n* = 29 controls, *n* = 29 HTG, *n* = 29 NTG, *n* = 27 PEXG). In all analyses, an FDR < 0.05 was considered significant.

### 2.3 mtDNA analysis

#### 2.3.1 DNA isolation and preparation

Buffy coat DNA from the study cohort subjects was isolated at the Department of Clinical Genetics at Maastricht University Medical Center. Isolations were performed with the QIAsymphony DSP Midi Kit (Qiagen) and the QIAsymphony platform (Qiagen) according to the manufacturer’s instructions in 7 batches of isolations containing randomly selected samples.

#### 2.3.2 mtDNA content measurement by real-time PCR

The number of mtDNA copies per cell, referred to as mtDNA content or mtDNA amount throughout the manuscript, was determined in DNA isolated from buffy coat by calculating the ratio of the mtDNA non-coding *D-loop* region to the nuclear housekeeping gene *B2M*. For that, a quantitative real-time PCR was performed on a LightCycler 480 (Roche) with the following setup: 10 min at 95 °C, 40 cycles of 15 sec at 95 °C and 1 min at 60 °C. A 12.5 µl reaction mix per gene containing 1x SensiMix™ SYBR^®^ & Fluorescein Kit (QT615-05, Bioline: Meridian Bioscience), 0.18 μM of forward (*D-loop* 5’ CATCTGGTTCCTACTTCAGGG 3’; *B2M* 5’ TGCTGTCTCCATGTTTGATGTATCT 3’) and reverse primer (*D-loop* 5’ TGAGTGGTTAATAGGGTGATAGA 3’; *B2M* 5’ TCTCTGCTCCCCACCTCTAAGT 3’), and ∼12.5 ng of DNA was added to each well of FrameStar^®^ 384 Well skirted PCR Plates (4ti-0381, 4titude). Both genes per participant were assessed in the same plate, in independent reactions, and in technical triplicate. The *D-loop* primers employed for the mtDNA copy number analysis have been previously verified using other mtDNA regions (*MT-ND1, MT-TL1*), and belong to a mtDNA region practically devoid of deletions.^47^

Raw fluorescence and melting curve data were exported from the LightCycler 480 (Roche) and loaded into the RDML-tools (https://www.gear-genomics.com) for melting curve, amplification curve and PCR efficiency corrected analysis with LinReg software ^48^. This LinReg software analyses the quality of the amplification and melting curves, and determines the efficiency of each PCR reaction, calculating the mean efficiency of each target per plate. Then it calculates the amount of each target copies (N0) using the Cq value and correcting it by the respective mean PCR efficiency of the target in that plate.

The ratio of PCR efficiency corrected initial copies of *D-loop* (mtDNA region) by *B2M* (nDNA gene) was performed to obtain the mtDNA content for each subject (N0 mtDNA / N0 nDNA). 6 balanced plates were performed with equal numbers of randomized subjects from each study group, with the respective paired treatment sample in the same plate (40 samples, 5 participants from each study group before and after treatment). 4 plates were run with samples that had to be repeated due to not having a proper triplicate (within <0.5 Cq) or flagged by the LinReg tool regarding the quality of the amplification or melting curve, in their respective balanced plates, to ensure the quality of the reaction of both targets per sample.

#### 2.3.3 mtDNA analysis and statistics

The mtDNA content per subject was analyzed using generalized linear mixed-effects modelling (Gamma distribution with an Inverse link and Gaussian random effects), including in the fixed effects the study groups (HTG, NTG, PEXG or Control), NAM treatment effect, and the interaction between the treatment and the groups. Several phenotypic covariates (sex, age, visual field index (%), diabetes, hypertension or specific medications (prostaglandin analogue, beta blocker, alfa agonist, carbonic anhydrase inhibitor, parasympaticomimetic drugs)) were added to the mentioned model in order to assess their possible relation to mtDNA content. In the random effects, the variable ‘Subject’ was included in order to pair subjects before and after treatment, and the variables “Run” and “Isolation” were included in order to correct for possible qPCR plate run and DNA batch of isolation experimental variability. Then, the inference criterion used for comparing the models is their ability to predict the observed data, i.e., models are compared directly through their minimized minus log-likelihood. When the numbers of parameters in models differ, they are penalized by adding the number of estimated parameters, a form of the Akaike information criterion (AIC). ^49^ The model under consideration was found to be better fitting if the AIC decreased compared to the previous model.

We identified that a generalized linear mixed-effects model with a Gamma distribution with an inverse link, and Gaussian random effects, fit the mtDNA content better than a linear model and generalized linear model with other distributions and links. The mtDNA content and Age were rescaled by dividing it by 100. For accurate interpretation and plotting of the model variables and its results, the estimates are first summed, then back-transformed by applying the inverse transformation due to the model’s link function, and finally multiplied by 100 to rescale the mtDNA content variable. Additionally, the estimates of Age are also multiplied by 100 due to their rescaling. All statistical analyses presented in regard to mtDNA content were performed using the freely available program R v4.3.1,^50^ and the ‘glmer’ function of the publicly available library ‘lme4’.^51^ All available 234 buffy coat samples (*n* = 30 controls, *n* = 30 HTG, *n* = 30 NTG, *n* = 29 PEXG; 119 pre-NAM / *n* = 30 controls, *n* = 27 HTG, *n* = 29 NTG, *n* = 29 PEXG; 115 post-NAM) were used for the generalized linear mixed-effects modelling.

## Results

### Cohort of study

The cohort has been described in Gustavsson et al., 2023.^46^

### Control and glaucoma subtypes have similar plasma metabolomes following oral Nicotinamide treatment

NAM’s long clinical history and good safety profile has enabled its rapid transition to clinical trials for glaucoma, but we lack an understanding of how NAM may affect patient metabolomes, and whether this differs between glaucoma subtypes. To determine this, we performed small-molecular weight high-resolution mass spectrometry of 114 subject plasma samples pre- and post-NAM treatment (see methods). We compared baseline (pre-NAM) plasma metabolomes (adjusting for age and sex) and identified minimal differences. Of the 94 identified metabolites that passed QC, only ethylmalonic acid was significantly different in glaucoma patients after adjustment for age and sex, demonstrating an elevation relative to control (Figure 1A). We next assessed the effect of 2 weeks of NAM on the plasma metabolome. Unsupervised hierarchical clustering of all 94 identified metabolites by all individuals pre- and post-NAM demonstrated that NAM treatment resulted in discrete metabolomes as evidenced by the clustering of two distinct groups which contained 113 pre-NAM (99%) and 2 post-NAM individuals (Group 1, “pre-NAM group”) and 112 post-NAM (98%) and 1 pre-NAM individuals (Group 2, “post-NAM group”; Figure 1B). This appeared to be driven largely by a cluster of 4 metabolites related to NAM (nicotinamide, 1-methylnicotinamide, 6-hydroxynicotinamide, N1-methyl-2-pyridone-5-carboxamide; Figure 1B). Individual pairwise comparison (pre vs. post treatment) confirmed this. Across all 4 groups, patients and controls, nicotinamide was significantly elevated (on the order of 20 to 170-fold increase) consistent with the high dose delivered (Figure 1C). We also detected a significant elevation of modified nicotinamide species (1-methylnicotinamide and 6-hydroxynicotinamide on a similar scale) and nicotinamide metabolites (N1-methyl-2-pyridone-5-carboxamide, 11 to 18-fold increase; Figure 1C). 5-methylcytosine was also significantly elevated in all groups, but the effect size was small (1.4 to 1.5-fold increase; Figure 1C). 4-pyridoxic acid was significantly decreased post-NAM treatment across all 4 subgroups (0.4-fold change) while betaine was significantly decreased in control and PEXG (Figure 1C). Sarcosine was significantly reduced in HTG and NTG, and hypoxanthine was significantly reduced in PEXG but the effect size of these was relatively small (0.7 to 0.8-fold change; Figure 1C). Supporting similar NAM responses across subgroups, construction of a linear model adjusting for age and sex demonstrated that only sarcosine responses were significantly different across groups, with significant reduction in HTG and NTG groups post NAM treatment (Figure 1D). Collectively, these results demonstrate that high dose NAM treatment increases NAM and NAM excretory/degradation products with minimal disruption to the plasma small-molecular weight metabolome after 2 weeks.

**Figure 1.**
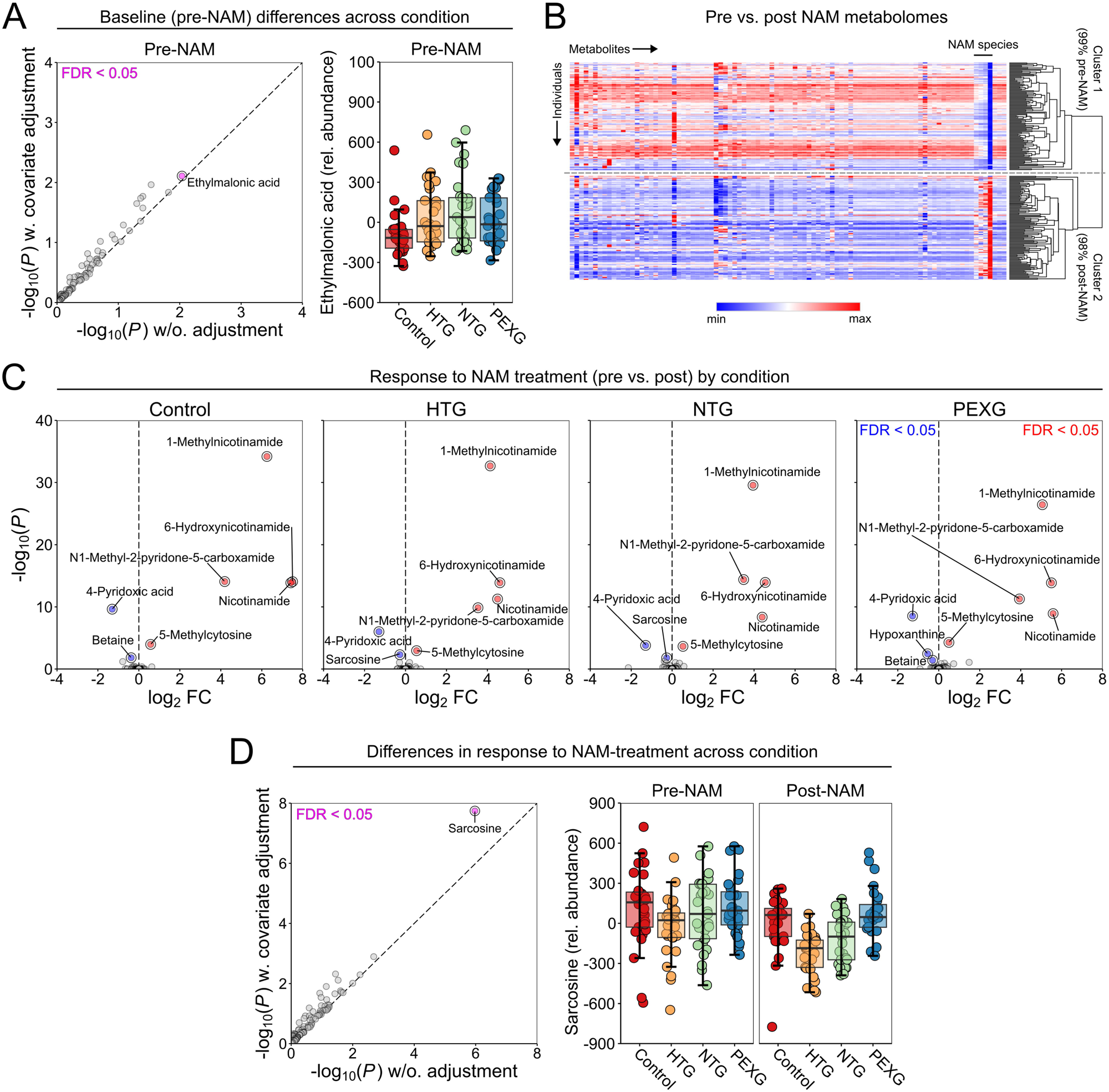
Control and glaucoma subtypes have similar plasma metabolomes following oral Nicotinamide treatment. **A)** Analysis by construction of a linear model with covariate adjustment (age, sex) demonstrated that baseline (pre-NAM) plasma metabolomes were similar across all subtypes compared to normal controls, with only ethylmalonic acid demonstrating a small yet significant relative increase in glaucoma. **B**) Unsupervised hierarchical clustering demonstrated that individual plasma samples clustered into pre- and post-NAM groups based on the metabolome alone, which appears to be largely driven by 4 NAM species metabolites. **C**) Individual comparisons of responses to NAM treatment (pre vs. post) demonstrated that all conditions respond similarly to high dose NAM treatment. Nicotinamide was significantly elevated across all conditions, and methylated NAM species were also detected suggesting active metabolism in the liver. In support of this, betaine (a methyl donor produced predominantly in the liver) was significantly reduced. **D**) Comparison of these responses by construction of a linear model with covariate adjustment (age, sex) demonstrated that sarcosine was the only metabolite that was significantly different across conditions. Sarcosine was significantly reduced in HTG and NTG patients but not in controls and PEXG.

### Control and HTG groups present a slight increase in buffy coat blood cell mtDNA content following oral NAM treatment

Next, we explored the possible effects of NAM supplementation on a mitochondrial marker measurable in blood, i.e. the amount of mtDNA in blood cells. For this, we assessed the mtDNA content in blood buffy coat DNA of glaucoma groups (HTG, NTG, PEXG) and healthy controls at baseline (pre-NAM) and after 2-week oral NAM treatment (post-NAM). This was done in DNA isolated from 234 buffy coats, including pre- and post-NAM treatment samples (see methods). The analysis was performed through a paired generalized linear mixed model with study groups, treatment effect and its interaction. We further assessed several phenotypic covariates and their possible relation with mtDNA content by adding them to the mentioned model. The best-fitting model was selected based on AIC (see methods). A final model was obtained with study groups, NAM treatment, along with sex and age as the relevant covariates that improved model fitting (Supplementary table 1). This model was simplified further by collapsing similar estimates, displaying the same statistical result (Supplementary table 2).

This analysis showed no differences in mtDNA content between the glaucoma groups and healthy controls at baseline (pre-NAM) (Figure 2A), nor did we observe differences between the groups post-NAM treatment (Figure 2B). Regarding the treatment response within the groups, we observed significant effects of the NAM treatment, as displayed by the main effects of the mixed model. The treatment effect differed in the NTG and PEXG groups from that of the other study groups, as indicated by the significant interactions of NTG and PEXG with NAM treatment. As depicted in Figure 2C, we observed the overall significant changes in mtDNA content within groups. The mtDNA content increased in controls and HTG after NAM treatment, whereas it did not in the NTG and PEXG study groups.

**Figure 2.**
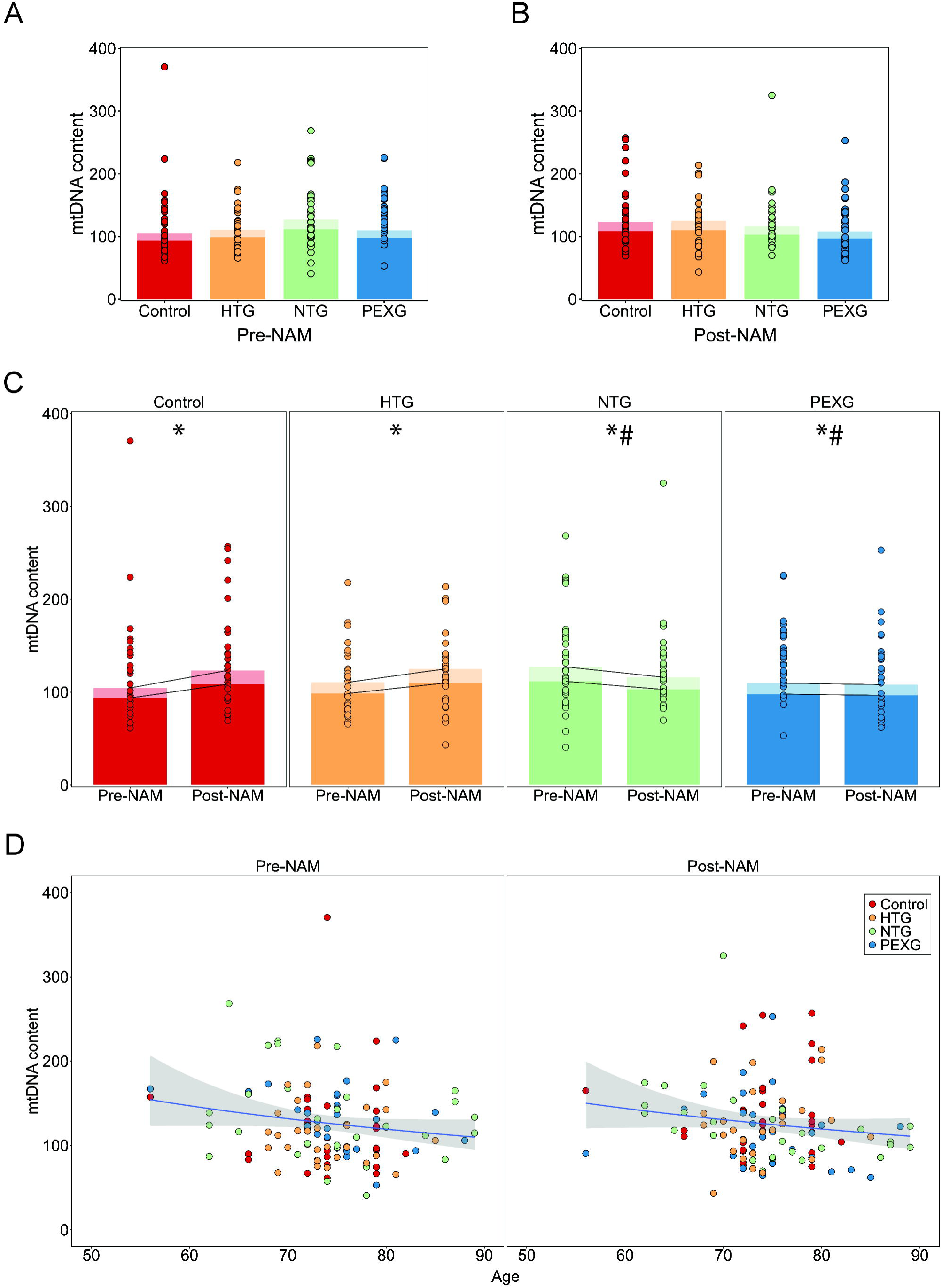
MtDNA content is comparable between glaucoma subtypes and the control group. A significant within-group slight increase is observed in the control and HTG groups following oral NAM treatment. **A, B)** Analysis through a paired generalized linear mixed-model with co-variates (age, sex) showed no differences between glaucoma groups and controls at baseline (pre-NAM) (**A**) and post-NAM treatment (**B**). **C)** The mixed-model displayed significant effects of NAM treatment in all study groups (*, main effects), and further that the NAM treatment effect in PEXG and NTG was significantly different (#, specific group interactions) from the effect observed in the control and HTG study groups. The mtDNA content increased in HTG and controls after NAM treatment whereas it did not in the NTG and PEXG study groups. **D)** A general decline of mtDNA with age was suggested by the mixed-model, occurring similarly pre- and post-NAM treatment. In **A**, **B** and **C**, the bright color in the bar represents the males and the fainter color the females, showing significantly lower mtDNA content in males than females in all the study groups pre- and post-NAM treatment, as indicated by the mixed-model main effects. Estimates from the mixed model (Supplementary table 1) are used for the construction of the bar plots (average of the calculated model back-transformed estimates) combined with dots representing the raw mtDNA content data. HTG: high tension Glaucoma; mtDNA: mitochondrial DNA; NTG: normal tension glaucoma; PEXG: pseudoexfoliative glaucoma.

In addition, we observed that mtDNA content differed between sexes, with males having a lower mtDNA content in all groups, as seen in the main effects of the mixed model (Figure 2A, 2B, 2C). No specific significant interaction with sex was found with any of the study groups (Supplementary figure 1). The model also suggested an overall trend for a decline of mtDNA content with age (Figure 2D). The model showed that this was a general trend, without any specific study group interaction (Supplementary figure 2).

No relation with disease severity, as measured by visual field index (best or only affected eye) was observed (Supplementary figure 3). No relation was also found with IOP levels (after IOP-related treatment), diabetes, or hypertension. These variables worsened the fit of the mixed model with study groups and treatment.

## Discussion

Targeting the bioenergetic decline in the RGCs by improving their capacity to generate and recycle NAD has shown to be a potential promising strategy in glaucoma. In the current study, we explored the effect of a 2 week NAM treatment in glaucoma and control subjects on the plasma metabolome and blood cell mtDNA content.

Metabolomic analysis of plasma revealed that control and glaucoma subtype metabolomes were largely similar, with an increase in ethylmalonic acid in glaucoma being the only significantly altered metabolite identified. Ethylmalonic acid is a branched fatty acid, the elevation of which is a strong biomarker of defects in short chain fatty acid β-oxidation. Previous studies have suggested an association between impairments in lipid metabolism with glaucoma.^52,53^ High levels of ethylmalonic acid (and its presence in urine) is typical of mutations to short chain acyl-CoA dehydrogenase (SCAD, *ACADS*),^54^ and ethylmalonyl CoA decarboxylase (*ECHDC1*).^55^ The presence of certain variants in these genes is linked to ethylmalonic encephalopathy syndrome but also other neurological symptoms, and conditions where flavin adenine dinucleotide (FAD) is low.^54–56^ When exposed to ethylmalonic acid, mitochondria demonstrate disruption to electron transport chain function, inhibition of mitochondrial creatine kinase,^57^) inhibition of succinate and malate transport,^58^ and other features of metabolic dysfunction.^59^ The increase in plasma ethylmalonic acid that we detected was modest, so it remains to be determined if this is sufficient to cause mitochondrial dysfunction or whether its elevation is high enough to mark mitochondrial dysfunction. Mild elevation of ethylmalonic acid is in line with, and perhaps another indicator of systemic mild metabolic deficiencies to mitochondria in glaucoma patients.^26,60^ Supporting this, pathway-based analysis of genetic variants has predicted alterations to Acetyl-CoA metabolism in POAG and NTG.^61^ While we were able to compare metabolomic profiles of control and glaucoma subtypes at baseline, the lack of major difference detected here is perhaps not surprising given the relatively small sample sizes of the subgroups. While previous metabolomic analyses have identified various differences between control and glaucoma patients, a recent meta-analysis of these highlights the difficulty in reproducibly identifying blood metabolome changes.^62^ Given that the primary aim of conducting the present metabolomic analysis was to identify NAM driven changes, future studies aimed at identifying underlying metabolic differences should aim for large group numbers.

To identify changes to the plasma metabolome induced by high dose NAM treatment, we analyzed plasma before and after treatment. By using paired samples, lower group sizes are more robust as individual variance is less impactful. We identified that NAM was significantly elevated in the plasma after 2 weeks of treatment as expected. Importantly, high dose NAM induced little change to the overall metabolome; 44% of changes were NAM and known NAM metabolites. 1-methylnicotinamide and 6-hydroxynicotinamide are produced by metabolism of NAM in the liver by cytochrome P450. Unaltered NAM is reabsorbed in the kidney and excretion requires conversion to 1-methylnicotinamide, but this can also be further metabolized to N1-methyl-2-pyridone-5-carboxamide by aldehyde oxidase.^63^ N1-methyl-2-pyridone-5-carboxamide is a reported uremic toxin, since its accumulation increases with chronic kidney disease (CKD). However, direct evidence of toxicity is conflicting and inconclusive.^63^ N1-methyl-2-pyridone-5-carboxamide is increased by over 100 fold in stage 2-3 CKD, 1000 fold in stage 4-5 CKD, and over 2000 fold in stage 5 CKD (hemodialysis patients).^63^ In comparison, we identified increases of 10 to 18-fold on average following NAM treatment, suggesting that potential kidney toxicity is unlikely. However, when patients with stage 5 CKD have been treated with high-dose NAM, severe increases of N1-methyl-2-pyridone-5-carboxamide have been demonstrated,^63^ supporting the need for careful use of NAM in patients with severe kidney disease. 4-pyridoxic acid was significantly decreased following NAM treatment (average 0.4-fold change). 4-pyridoxic acid is a product of vitamin B_6_ catabolism by aldehyde oxidase,^64^ and so this reduction of 4-pyridoxic acid is likely explained by competition for the enzyme by conversion of 1-methylnicotinamide to N1-methyl-2-pyridone-5-carboxamide. We also identified a reduction in plasma betaine, an important methyl donor predominantly found in the liver, which may be a consequence of the need for methylation of NAM to 1-methylnicotinamide. 1-methylnicotinamide, although predominantly considered a clearance product, has demonstrated positive vascular effects including anti-thrombotic, anti-inflammatory, and antioxidant effects.^65^ Hypoxanthine was significantly reduced only in PEXG following NAM treatment, but this was no longer significant when adjusting for age and sex. Sarcosine, however, remained significantly reduced after adjustment. Sarcosine was significantly lower in HTG and NTG patients. Sarcosine (methylglycine) can be generated from glycine (and vice versa) within mitochondria or from dietary choline (via betaine). Loss of sarcosine can be indicative of a folate deficiency since tetrahydrofolic acid is required for the inter-conversion of sarcosine and glycine.^66^ This glycine–sarcosine cycle is an important regulator of the ratio of the universal methyl donor S-adenosylmethionine (SAM) to its demethylated form S-adenosylhomocysteine (SAH), an indicator of methylating capacity.^67^ Reduced sarcosine in HTG and NTG patients, but not controls, may be an indicator of reduced methylating capacity. We have previously demonstrated that the methyl cycle and one-carbon metabolism is disrupted in the retina in glaucoma models and that POAG patients harbor some indicators of disrupted mitochondrial one carbon metabolism.^68^ This suggests that there may be some utility in combining methyl boosting supplements with NAM long-term in some patient subtypes.

The mtDNA analysis of blood buffy coat DNA showed no difference in blood cell mtDNA amount between control and glaucoma subtypes in the current study. Previous studies in whole blood or buffy coat have found differences in mtDNA levels of glaucoma vs. controls, such as increased levels in a Japanese population with mixed NTG and HTG, and decreased levels in a Dutch population.^69,70^ In isolated lymphocytes, no differences between NTG and Controls were detected in a UK population.^71^ This discrepancy in outcomes might be attributed to population differences, for instance at a genetic background level,^72,73^ sampling/methodological effects, or the heterogeneity of glaucoma (disease progression/severity at time of sampling), but does not exclude mtDNA as a relevant biomarker.

We observed that blood cell mtDNA content increased in the control and HTG groups after NAM treatment, whereas this was not observed in the NTG and PEXG groups. The observed increase in both controls and HTG groups was modest (∼12% and ∼17% respectively). Previous studies *in vitro* have reported an increase in mtDNA amount and mtDNA transcripts in osteoblasts and fibroblasts following NAM supplementation.^74,75^ Supplementation *in vitro* with nicotinamide mononucleotide (NMN), the first product of NAM processing in the salvage pathway, led to an increase in mtDNA copy number in HEK293, which is suggested to occur through the activation and enhancement of mtDNA replication.^76^ In *in vivo* studies, other NAD metabolites from the salvage pathway, such as NR, have been shown to improve mitochondrial biogenesis and increase mtDNA abundance in the muscle tissue of mice. An increased expression, at mRNA level, for genes encoding proteins related to these processes has also been observed, e.g., PGC-1α, TFAM, CS.^77–79^ These effects may reflect enhanced activity of sirtuins, NAD+-dependent histone deacetylases, which increase their activity in response to elevated NAD+ availability. Interestingly, sirtuins such as SIRT1 have been studied in relation to ocular aging, and the NAM neuroprotective effect has been suggested to be related to the NAD+/SIRT1 axis.^80–82^ The short nature of the treatment in this study could be the reason that we only found a slight increase, and that this was not observed in NTG and PEXG. However, we cannot exclude that differences in disease phenotype could lead to varying treatment responses.

### Limitations

We detected limited baseline metabolomic differences between glaucoma patient subtypes and controls which may relate to the relatively small samples sizes (∼30 / condition). Larger sample sizes that better reflect the heterogeneity of glaucoma patients and repeat measures which reduce the likelihood of snapshot findings may be required to identify more disease-related metabolic differences in patient blood. Comparison of pre- and post-NAM-treatment effects was more robust given that we could conduct pairwise analysis, but the treatment duration was relatively short-term and so we cannot preclude larger changes with a longer treatment course.

We used buffy coat DNA for assessing the blood cell mtDNA content. Since buffy coat contains all types of leukocytes, the measurements of this heterogeneous cell population may be more variable as compared to measurements of isolated and purified peripheral blood mononuclear cells (PBMCs). Additionally, buffy coat may contain a fraction of platelets, which contain mtDNA but lack nuclei (and thus present potential source of variation for the measured ratio of mtDNA content). In order to minimize technical variability, we took care to conduct blood draw, sample processing, buffy coat collection and DNA isolation systematically and uniformly for all participants. The study groups were also age- and sex-matched.

## Conclusion

In conclusion, 2 weeks NAM supplementation resulted in a largely consistent plasma metabolome response across glaucoma subtypes and healthy controls, which predominantly reflected increased NAM metabolites and intermediates, with minimal effects on the wider metabolome. NAM supplementation also led to a modest increase in blood mtDNA amount in HTG and controls. These results indicate an overall safe and positive response to the treatment observed under short-term supplementation and a limited cohort size. Ongoing long-term and larger cohort clinical studies (**NCT05275738**) will be essential to clarify further systemic effects of NAM treatment and to strengthen the evidence for its neuroprotective therapeutic potential in glaucoma.

## Declarations

### Ethics approval

The study was approved by the Swedish Ethical Review Authority (2020-01525, 2021-01036, 2021-03745, and 2022-04851).

### Consent for publication

Not applicable.

### Availability of data and materials

All data generated or analyzed during this study will be included in the final peer-reviewed published article.

### Author’s contributions

AVG – performed experiments, analyzed data, wrote the manuscript.

JRT – performed experiments, analyzed data, wrote the manuscript.

SG – performed experiments, analyzed data, wrote the manuscript.

BJB – provided supervision, wrote the manuscript.

PJL – provided supervision, wrote the manuscript.

CABW – provided supervision, wrote the manuscript.

HJMS – provided supervision, wrote the manuscript.

GJ – provided supervision, conceived and designed experiments, wrote the manuscript.

TGMFG – provided supervision, conceived and designed experiments, wrote the manuscript

PAW – provided supervision, conceived and designed experiments, wrote the manuscript.

All authors read and approved the final manuscript.

## Acknowledgements

The authors would like to thank St. Erik Eye Hospital for financial support for research spaceand facilities. Swedish Metabolomics Centre, Umeå, Sweden (www.swedishmetabolomicscentre.se) is acknowledged for metabolic profiling by LCMS. The authors would like to thank the following foundations: Glaucoomfonds and Oogfonds, which contributed through UitZicht to this research. The funding organizations had no role in the design or conduct of this research. They provided unrestricted grants.

