## Supplementary Figures for "Short-term high-dose nicotinamide treatment across glaucoma subtypes reveals increased mtDNA content and minimal metabolomic change in blood"

**Supplementary figure 1.** Relation between mtDNA content and sex in the different glaucoma subtypes and controls at baseline (pre-NAM) and post-NAM treatment. HTG: high tension Glaucoma; NAM: Nicotinamide; NTG: normal tension glaucoma; PEXG: pseudoexfoliative glaucoma.

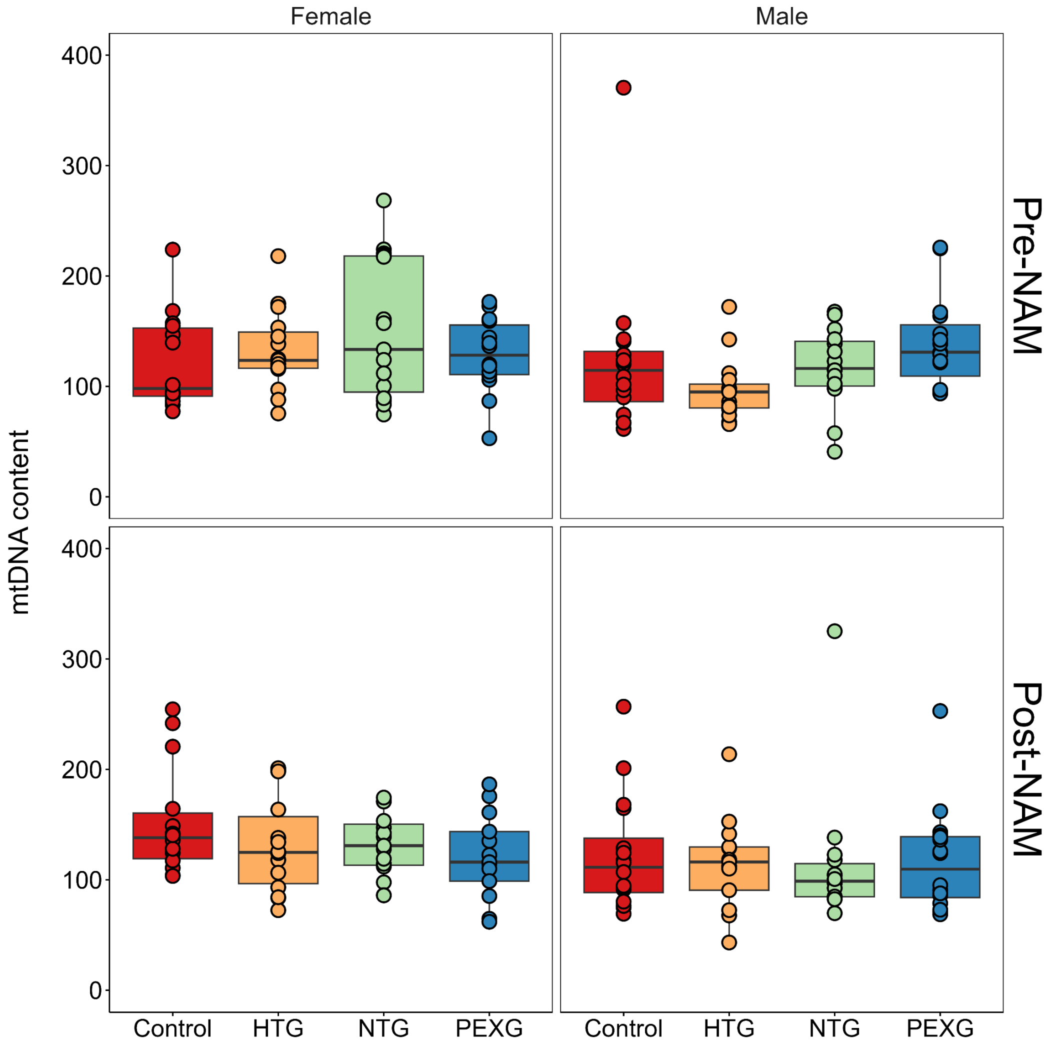

**Supplementary figure 2.** Relation between mtDNA content and age in the different glaucoma subtypes and controls at baseline (pre-NAM) and post-NAM treatment. HTG: high tension Glaucoma; NAM: Nicotinamide; NTG: normal tension glaucoma; PEXG: pseudoexfoliative glaucoma.

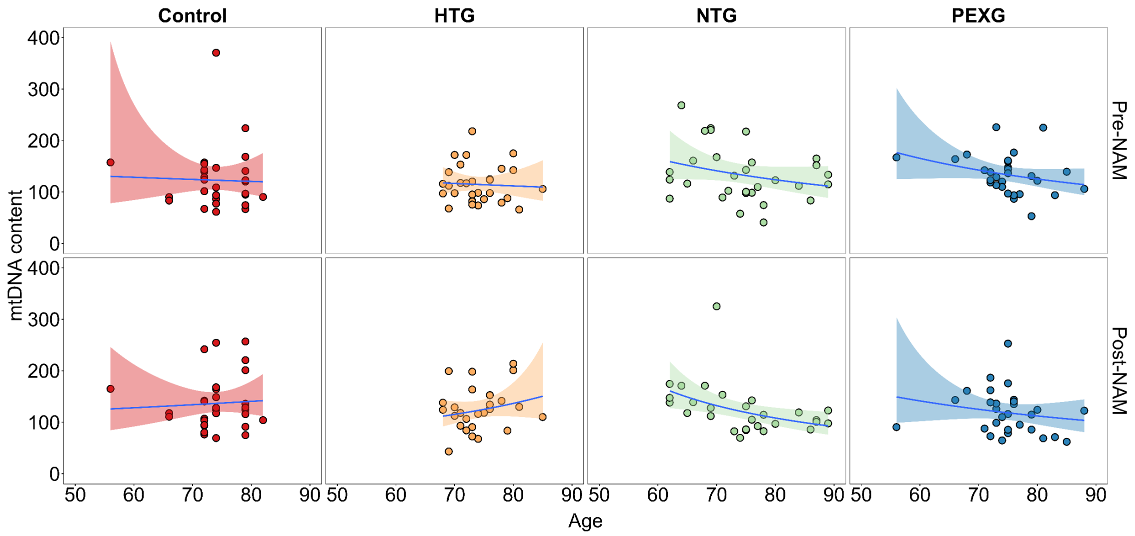


**Supplementary figure 3.** Relation between mtDNA content and visual field index (%) (best or only affected eye) in the different glaucoma subtypes and controls at baseline (pre-NAM) pre and post-NAM treatment. HTG: high tension Glaucoma; NAM: Nicotinamide; NTG: normal tension glaucoma; PEXG: pseudoexfoliative glaucoma.

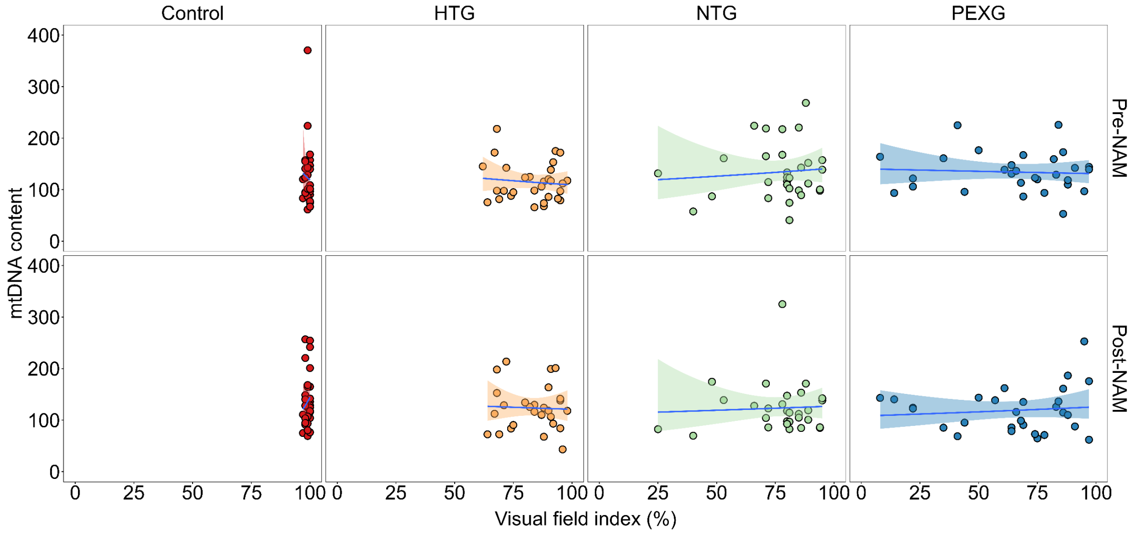
