## Supplementary Tables for "Short-term high-dose nicotinamide treatment across glaucoma subtypes reveals increased mtDNA content and minimal metabolomic change in blood"

**Supplementary table 1.** Final generalized linear mixed model.

| | mtDNA content | | |
|---|---|---|---|
| *Predictors* | *Estimates* | *Std. error* | *p-value* |
| Controls (Intercept) | 0.40433 | 0.32056 | 0.207 |
| HTG | -0.05228 | 0.08614 | 0.544 |
| NTG | -0.17020 | 0.08809 | 0.053 |
| PEXG | -0.04456 | 0.07743 | 0.565 |
| NAM Treatment | -0.14561 | 0.05395 | **0.007** |
| Sex (Male) | 0.11021 | 0.04990 | **0.027** |
| Age | 0.74191 | 0.41573 | 0.074 |
| HTG : NAM Treatment | 0.04137 | 0.07433 | 0.578 |
| NTG : NAM Treatment | 0.22111 | 0.07229 | **0.002** |
| PEXG : NAM Treatment | 0.15847 | 0.07573 | **0.036** |
| **Random Effects** | | | |
| $\sigma^2$ | 0.07226 | | |
| $\tau_{Subject}$ | 0.02503 | | |
| $\tau_{Plate\ Run}$ | 0.00349 | | |
| $\tau_{DNA\ Isolation}$ | 0.00376 | | |
| $N_{Plate\ run}$ | 10 | | |
| $N_{Subject}$ | 120 | | |
| $N_{DNA\ Isolation}$ | 7 | | |
| Observations | 234 | | |
| Marginal $R^2$ / Conditional $R^2$ | 0.080 / 0.364 | | |
| AIC | 192.681 | | |

Final model (Gamma distribution with an Inverse link and Gaussian random effects) including age and sex which improved the fitting of the base model with study groups and treatment. The model fits better than the null model (without variables, AIC = 193.41). MtDNA content and age were rescaled by dividing them by 100. Estimates have to be back-transformed by doing the inverse, due to the link of the model (See Methods, statistical analysis). The back-transformed estimates of this model were used to construct the plots in Figure 2 (A,B and C). P-values < 0.05 are marked in bold. HTG: high tension glaucoma; N: number of levels of groups in the random effects; NTG: normal tension glaucoma; PEXG: pseudoexfoliative glaucoma; std: standard; $\tau$: variance of the random effects set intercepts; $\sigma^2$: residual variance; (:) indicates interaction between two variables. NAM: nicotinamide

**Supplementary table 2.** Simplified final generalized linear mixed model with collapsed similar estimates.

| | mtDNA content | |
|---|---|---|
| *Predictors* | *Estimates* | *Std. error* |
| Controls + HTG (Intercept) | 0.39845 | 0.31778 |
| NTG + PEXG | -0.08320 | 0.05468 |
| NAM Treatment | -0.12121 | 0.04515 |
| Sex (Male) | 0.11228 | 0.04997 |
| Age | 0.70524 | 0.41549 |
| (NTG + PEXG) : NAM Treatment | 0.17247 | 0.05881 |
| **Random Effects** | | |
| $\sigma^2$ | 0.07321 | |
| $\tau_{Subject}$ | 0.02524 | |
| $\tau_{Plate\ Run}$ | 0.00329 | |
| $\tau_{DNA\ Isolation}$ | 0.00304 | |
| $N_{Plate\ run}$ | 10 | |
| $N_{Subject}$ | 120 | |
| $N_{DNA\ Isolation}$ | 7 | |
| Observations | 234 | |
| Marginal $R^2$ / Conditional $R^2$ | 0.066 / 0.347 | |
| AIC | 186.750 | |

Simplified final model (Gamma distribution with an Inverse link and Gaussian random effects) by collapsing variables in the final model that have similar model estimates improving the model further. The model fits better than the null model (without variables, AIC = 193.41). MtDNA content and age were rescaled by dividing them by 100. Estimates have to be back-transformed by doing the inverse, due to the link of the model (See Methods, statistical analysis). HTG: high tension glaucoma; N: number of levels of groups in the random effects; NTG: normal tension glaucoma; PEXG: pseudoexfoliative glaucoma; std: standard; $\tau$: variance of the random effects set intercepts; $\sigma^2$: residual variance; (:) indicates interaction between two variables. NAM: Nicotinamide.
